# Microglial function moderates the relation between depression risk factors and depression outcomes across the life course in females

**DOI:** 10.1101/2023.02.18.23286124

**Authors:** Eamon Fitzgerald, Irina Pokhvisneva, Sachin Patel, Shi Yu Chan, Ai Peng Tan, Helen Chen, Patricia Pelufo Silveira, Michael J Meaney

**Affiliations:** Ludmer Centre for Neuroinformatics and Mental Health, McGill University, Canada; Douglas Mental Health University Institute, Department of Psychiatry, McGill University, Canada; Translational Neuroscience Program, Singapore Institute for Clinical Sciences, Singapore; Yong Loo Lin School of Medicine, National University of Singapore, Singapore; Department of Diagnostic Imaging, National University Health System, Singapore; Brain – Body Initiative, Agency for Science, Technology & Research (A*STAR), Singapore; Department of Psychological Medicine, KK Women’s and Children’s Hospital, Singapore; Duke-National University of Singapore, Singapore

**Keywords:** Depression, microglia, sex differences, genetics, psychiatric disorders, risk factors

## Abstract

**Background:** Depression has an enormous socio-economic burden and is twice as common in women compared to men. Microglia are exceptionally responsive to environmental stimuli and their phenotype differs substantially by sex. We hypothesized microglial function would moderate the relation between depression risk factors and depressive outcomes in a sex-specific manner.

**Methods:** We used expression quantitative trait loci and single nucleus RNA-sequencing resources to generate polygenic scores (PGS) representative of individual variation in microglial function in the fetal (GUSTO; N=239-315, and ALSPAC; N=928-1461) and adult periods (UK Biobank; N=54753-72682). We stratified our analyses by sex and tested the interaction effects of these PGS with prenatal maternal depression symptoms and adult stressors, well-characterized depression risk factors. We used internalizing (early childhood) or depressive symptoms (late childhood and adulthood) as outcomes.

**Results:** The fetal microglia PGS moderated the association between maternal prenatal depressive symptoms and female offspring internalizing symptoms at 4 (GUSTO; beta=-0.25, 95%CI -0.44 to - 0.06, P=0.008) and 7 years (GUSTO; beta=-0.16, 95%CI -0.318 to -0.008, P=0.04), and depressive symptoms at 8.5-10 years (GUSTO; beta = -0.15, 95%CI = -0.25 to -0.03, P= 0.01) and 24 years (ALSPAC; beta=0.1, 95%CI 0.008 to 0.19, P=0.03). The adult microglial PGS moderated the relation between BMI (UK Biobank; beta=0.001, 95%CI 0.0009 to 0.003, P=7.74E-6) and financial insecurity (UK Biobank; beta=0.001, 95%CI 0.005 to 0.015, P=2E-4) with depressive symptoms in females. There were no significant interactions in males.

**Conclusion:** Our results illustrate an important role for microglial function in the conferral of sex-dependent depression risk.

## Introduction

Depression is a debilitating disorder with profound socio-economic consequences. In 2019 depression was the second leading contributor to years lived with disability worldwide (1). This burden has increased considerably during the COVID-19 pandemic (2). The economic burden of depression is as dramatic, with costs estimated at $210.5 billion in the United States alone in 2010 (3). These costs are disproportionately shouldered by women, with estimates of depression incidence indicating a 2-fold higher rate of depression in women over men (1,4,5). Somewhat curiously the mechanisms behind these sex-differences are not well studied despite evidence for sex-specific pathways (6,7).

Even the highest estimates of overall depression heritability fall well below 50% (8), illustrating a strong role of the environment in depression risk. Epidemiological studies consistently show strong links between depression and adverse experiences in both early life and adulthood, which persist across cultures, sex, gender and ancestry (9–11). These studies have highlighted many exposures that act to increase depression risk, including perinatal exposures (e.g. exposure to prenatal maternal depression (12–14)), adult exposures (e.g. financial insecurity (15)) and physiological measures (e.g. BMI (11,16)).

How these diverse environmental exposures translate to increased depression risk and why this occurs with such sex-specificity is largely unclear. Much of the research in this area has been performed using animal models where many elegant studies have identified neuronal plasticity as an important regulator of the behavioural response to chronic stress and the subsequent therapeutic attenuation of these behaviours (17). Glial cells comprise the majority of cells in the brain and have a critical role in many aspects of neuronal plasticity (18), but their role in the behavioural response to adverse environments has been vastly understudied. Even fewer studies have investigated the role of glia in human depression risk, but several streams of evidence suggest a possible contribution, particularly with respect to sex-dependent depression risk.

Microglia are one such glial cell that satisfies several criteria to confer sex-dependent depression risk. First, they are exceptionally responsive to the environment (18), with well-characterized homeostatic roles in regulating synapses (19), neuronal function (20) and myelination (21). Second, in model systems microglia display sexual dimorphisms in their morphology, distribution, electrophysiology, transcriptome and proteome (22–25). Although less studied, transcriptomic differences between male and female microglia have also been described in humans (26). Third, alterations in microglia drive the behavioural response to both early life and adult stresses in non-human, animal models (27–30). Fourth, changes in microglia have been described in humans with depression using histological, *in vivo* imaging and transcriptomic approaches (31–34). Interestingly, these transcriptional changes existed largely for genes with homeostatic functions (33,34). Together these studies led us to hypothesize that microglia would moderate the relation between environmental risk factors and depression outcomes, and that this associations would occur in a sex-dependent fashion.

We tested this hypothesis using a functional genomics approach in 3 human cohorts (2 birth cohorts, 1 adult cohort). We leveraged single cell RNA sequencing (scRNA-seq) and expression quantitative trait loci (eQTL) resources to generate polygenic scores (PGS) representative of individual variation in microglial function. We generated these scores for individuals in the Growing Up in Singapore Towards healthy Outcomes (GUSTO), Avon Longitudinal Study of Parents and Children (ALSPAC) and UK Biobank cohorts. We then tested the moderation effects of these PGS on the relation between depression risk factors and depression outcomes. In GUSTO and ALSPAC we created a microglial PGS using scRNA-seq and eQTL data from the mid-gestational human fetal cortex. In these cohorts we found the microglial PGS moderated the relation between prenatal maternal depressive symptoms and offspring depressive outcomes only in females. We then created an adult microglial PGS, using scRNA-seq and eQTL data from non-diseased human adult anterior cingulate cortex tissue, in the UK Biobank. We show that this adult microglial PGS moderated the relation between depression risk factors (BMI and financial stress) and specific depression symptoms, but again only in females.

Together our results describe a remarkable specificity for functional variants associated with microglial enriched genes to moderate the relation between environmental risk factors and depressive outcomes in females.

## Methods and materials

### Cohorts

GUSTO is a Singapore-based longitudinal birth cohort that recruited pregnant women, older than 18 years of age, from two Singaporean birthing hospitals with data collection of mothers and offspring (35). Ethical approval for the GUSTO study was attained from the National Healthcare Group Domain Specific Review Board (NHG DSRB; D/2009/021 and B/2014/00411) and the SingHealth Centralized Institutional Review Board (CIRB; D/2018/2767 and A/2019/2406). All participating mothers provided informed consent. Detailed information about the GUSTO study can be found at https://gustodatavault.sg/.

ALSPAC is a UK-based longitudinal birth cohort from the southwest of England. Pregnant women within the Avon area, with estimated delivery dates between April 1^st^, 1991 and December 31^st^, 1992 were invited to participate (36–38). There were 14,541 pregnancies initially enrolled in the study, resulting in 14,676 foetuses from 14,062 live births, of which 13,988 children were alive at 1 year of age. ALSPAC data were collected and managed using REDCap electronic data capture tools hosted at the University of Bristol. REDCap (Research Electronic Data Capture) is a secure, web-based software platform designed to support data capture for research studies. (39). Ethical approval for the ALSPAC study was granted by the ALSPAC Ethics and Law Committee and local research ethics committees (full list available at http://www.bristol.ac.uk/alspac/researchers/research-ethics/). Informed consent was obtained from participants in line with these approved ethical guidelines. Full details of ALSPAC data are searchable at: http://www.bristol.ac.uk/alspac/researchers/our-data/.

The UK Biobank is a UK-based population cohort of adults. Ethical approval for this cohort was obtained from the Northwest Multicentre Research Ethics Committee (REC reference 11/NW/0382), the National Information Governance Board for Health and Social Care and the Community Health Index Advisory Group. All participants provided informed written consent before data collection. The research in this manuscript was conducted under application number 41975.

Throughout this study, sex was defined genetically. Analysis in the GUSTO and ALSPAC cohorts were confined to term, singleton births. See **Supplementary Table 1** for a description of cohort demographics. See the Supplementary methods for a full description of outcome and genotype data collection and analysis.

### Statistical analysis

All analyses, unless otherwise stated, were conducted with R v4.1.1 (51) and RStudio v1.4.1717 (52). Linear and logistic regression analyses were performed using the lm() or glm() functions. We used the influence.measures() function in R to identify samples with undue influence on the regression results. Any samples with excessive influence were removed and a new regression model was calculated. This was not done in the UK Biobank analysis due to the large sample size. Specific covariates and sample size for each comparison are mentioned in the results. If a significant interaction was found in the regression model, a simple slopes analysis was performed using the Interactions package (53). PGS were treated as continuous variables in all analyses. Where appropriate correction for multiple comparisons was done using the Benjamini-Hochberg method (false discovery rate; FDR). Plotting was done using either the Interactions package or ggplot2 (54). An alpha level of 0.05 for statistical significance was used throughout.

## Results

### A microglial polygenic score moderates the relation between prenatal maternal depression and child internalizing and depressive symptoms in females of the GUSTO cohort

Prenatal maternal depression is a major risk factor for offspring depression (12,55). We hypothesized that microglial function would moderate this relation in a sex-specific manner. To test this hypothesis, we identified microglial enriched genes from a scRNA-seq study of the mid-gestational fetal cortex (gestational week 17-18; Polioudakis et al., 2019; **Supplementary Table 2**) and used eQTL data from Walker et al (an eQTL study of mid-gestational cortex) to generate a PGS representative of individual variation in microglial-enriched gene expression for children of the GUSTO cohort (see methods section).

We used multivariate regression stratified by males and females, to estimate the interaction between the microglial PGS and EPDS using the internalizing scale of the Child Behaviour Checklist (CBCL) as the outcome. If a significant interaction was found (P<0.05), a simple slopes analysis was used to characterize the specific associations. The first 10 genetic principal components and child gestational age at birth were included as covariates. We found a significant interaction between the microglial PGS and prenatal maternal EPDS in females using child internalizing symptoms at 4 and 7 years of age as the outcomes (**Figure 1A and 1B** and **Supplementary Table 3**; 4 years-beta=-0.25, 95%CI -0.44 to -0.07, P=0.008; 7 years-beta=-0.16, 95%CI -0.318 to -0.008, P=0.04). This relation was such that a lower PGS in the child increased risk of internalizing symptoms when exposed to a higher prenatal maternal EPDS. Significant interaction effects were unaffected when a polygenic risk score (PRS) for depression was used as a covariate in the analysis (**Supplementary Table 4**). Identical analyses for males did not identify any significant interaction effects between the microglia PGS and prenatal EPDS (**Supplementary Table 5**). A similar approach using data from other cell-types (outer radial glia, oligodendrocyte precursor cells and excitatory neurons; **Supplementary Table 6 and Supplementary Table 7**) revealed no significant moderation in females suggesting this association was specific to microglia. We also found no significant interaction effects when using externalizing symptoms as the outcome in females (4 years- beta=-0.12, 95%CI -0.29 to 0.05, P=0.16; 7 years-beta=-0.08, 95%CI -0.25 to 0.08, P=0.3; **Supplementary Table 3**), suggesting our results are specific for females, internalizing symptoms, and microglia.

**Figure 1.**
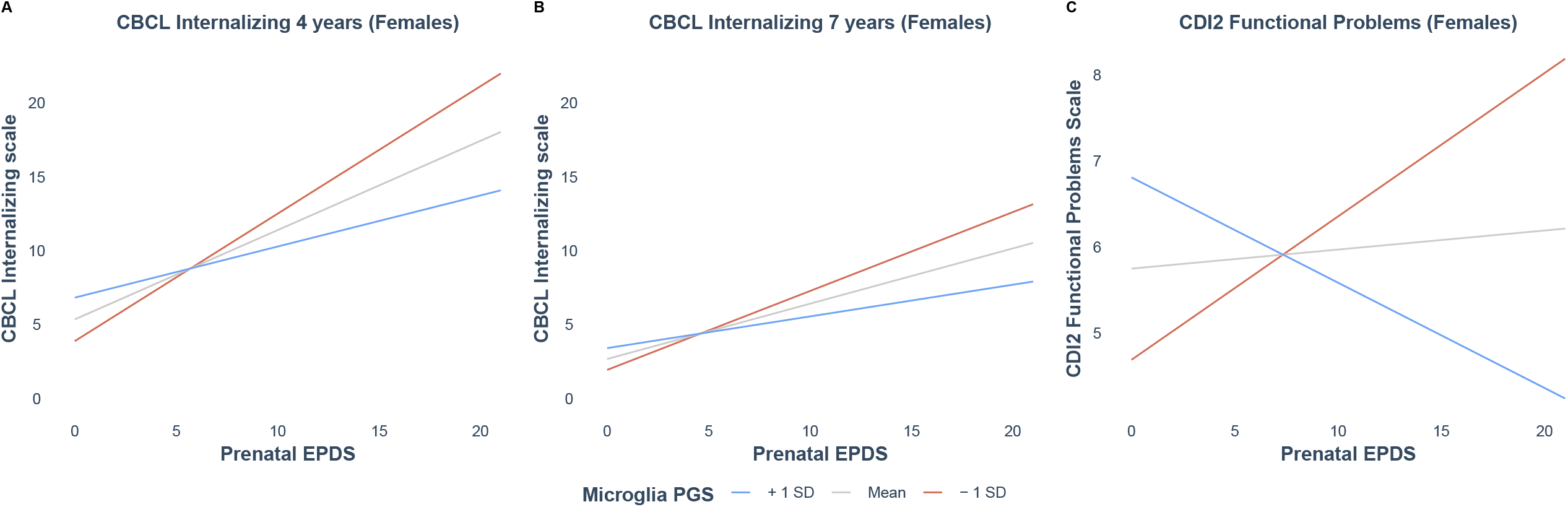
A prenatal microglial PGS moderates the relationship between prenatal maternal depressive symptoms and offspring internalizing or depressive outcomes during childhood in females of the GUSTO cohort. All panels represent the results from a simple slopes analysis. For all panels, the relationship between maternal prenatal EPDS at 26 weeks post-conception (x-axis) and offspring outcome (y-axis) is stratified by higher (+1 SD; blue line), lower (-1 SD; red line) or mean (grey line) microglial PGS in offspring. A) A significant interaction effect was seen for the CBCL internalizing score at 4 years in GUSTO (N= 313; lower PGS-slope= 0.86, SE=0.14, P=7.18E-10; higher PGS-slope=0.35, SE=0.14, P=0.01). B) A significant moderation was seen for CBCL internalizing score at 7 years in GUSTO (N= 239; lower PGS-slope= 0.53, SE=0.11, P=3.31E-6; higher PGS-slope=0.21, SE=0.11, P=0.05). C) A significant moderation was seen for the Functional Problems Scale of the CDI2 at 8.5-10 years in GUSTO (N= 301; lower PGS-slope=0.17, SE=0.08, P=0.04; higher PGS-slope=-0.12, SE=0.08, P=0.11).

We then used a similar approach to assess the interaction between maternal EPDS and the microglial PGS using the Child Depression Inventory 2 (CDI2) as the outcome. The CDI2 is a child-reported questionnaire that specifically assesses depressive symptoms and was administered once between 8.5 to 10 years of age in GUSTO. Age at administration was included as an additional covariate in this analysis. There was a significant interaction effect between the microglia PGS and EPDS in females when the CDI2 total score was used as the outcome (beta=-0.22, 95%CI -0.43 to - 0.02, P=0.03), but a simple slopes analysis did not identify a significant difference for either the high or low PGS (high PGS-slope = -0.17, SE=0.14, P=0.23; low PGS-slope=0.28, SE=0.15, P=0.06; **Supplementary Table 3**). We then investigated the scales comprising the total CDI2 score and identified a significant interaction when the Functional Problems scale (measures dysfunction in social relationships and one’s interpretation of their academic performance) was used as the outcome (beta = -0.14, 95%CI = -0.25 to -0.03, P= 0.01). Simple slopes analysis then showed a significant association for the lower PGS (high PGS-slope=-0.12, SE=0.08, P=0.11; low PGS-slope=0.17, SE=0.08, P=0.04; **Figure 1C**). This result yields an equivalent interpretation to the CBCL analysis, a lower microglial PGS is associated with increased risk of depressive symptoms in childhood when exposed to a mother with a high prenatal EPDS. This interaction effect remained significant when a PRS for depression was included as a covariate in the analysis (**Supplementary Table 4**). There were no significant interaction effects with anxiety outcomes (Multidimensional Anxiety Scale for Children 2; MASC2; **Supplementary Table 3**), in males (**Supplementary Table 5)** or with similar PGS from other fetal cell-types (**Supplementary Table 6 and 7**). Again, these results suggest specificity for females, microglia, and depressive outcomes.

### A microglia PGS moderates the association of prenatal maternal EPDS on adult depressive outcomes in the ALSPAC cohort

The trajectory of depressive symptoms from childhood to adulthood is complex (57). We therefore, wanted to assess whether the microglial PGS associations we described in childhood persist into adulthood. To this end we used identical criteria to generate a microglia PGS in the UK birth cohort, ALSPAC, and explored the interaction effect with prenatal maternal EPDS using depressive symptoms at 24 years as the outcome. We note that the period covered by the GUSTO and ALSPAC data sets includes the peak age of onset for depression. We again stratified analyses by sex. Depressive symptoms were assessed using the Clinical Interview Schedule-Revised edition (CIS-R), a validated proxy for a clinical interview in the assessment of mental health outcomes. We used the first 10 genetic principal components and gestational age at birth as covariates in the analysis. We found significant interactions between the microglial PGS and EPDS in females when mild, moderate or severe depression was used as the outcome (mild-beta=0.04, 95%CI 0.007 to 0.08, P=0.02; moderate-beta=0.05, 95%CI 0.008 to 0.09, P=0.02, severe-beta=0.1, 95%CI 0.008 to 0.19, P=0.03). Simple slopes analysis showed a significant associations for the higher PGS, which was consistent across outcomes (**Figure 2A-2C and Supplementary Table 8**). There were no significant interaction effects when equivalent analyses were conducted in males (**Supplementary Table 10**). The interpretation of these results is that a higher PGS in females is associated with increased risk of adult depression when exposed to a high prenatal maternal EPDS. This contrasts with childhood internalizing and depressive symptoms, where the lower PGS was associated with increased risk. The results for mild and moderate depressive outcomes remained when a depression PRS was used as a covariate, but the interaction with severe depression was attenuated (beta=0.07, 95%CI -0.03 to 0.16, P=0.19; **Supplementary Table 9**). There were no significant interaction effects seen when using PGS based on different fetal cell-types (**Supplementary Table 8**) or when anxiety was used as the outcome (**Supplementary Table 8**). Similar to our results in GUSTO, this suggests specificity for females, microglia and depressive symptoms.

**Figure 2.**
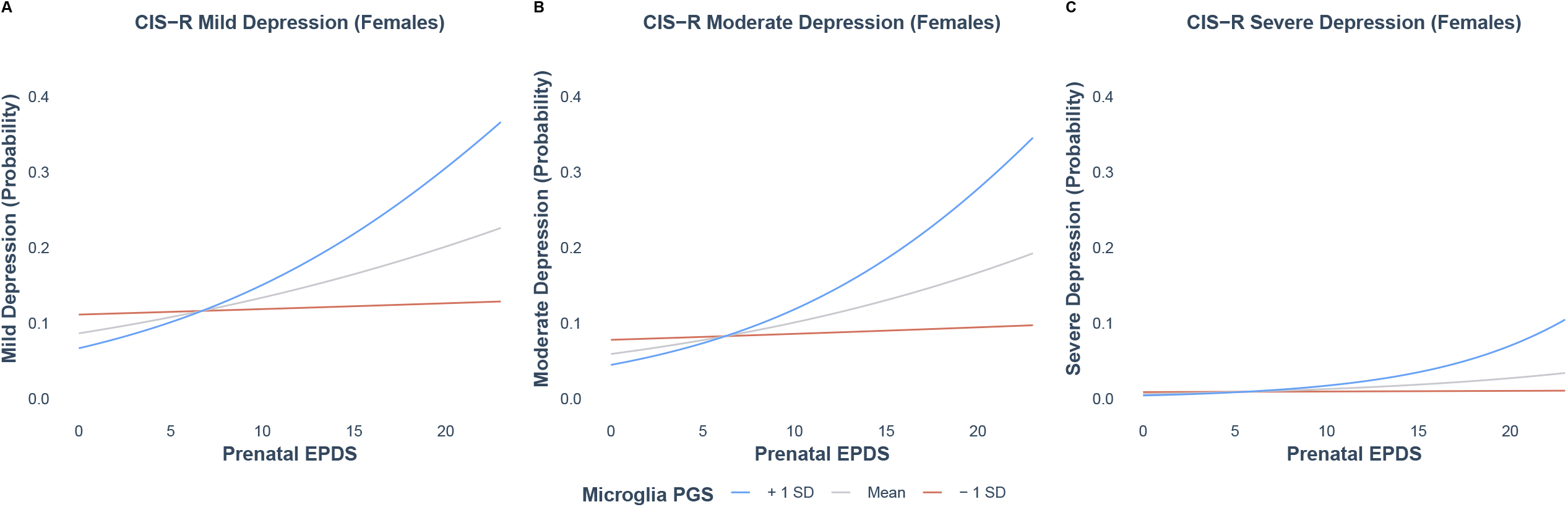
A prenatal microglia PGS moderates the relationship between prenatal maternal EPDS and offspring depressive outcomes at 24 years in females of the ALSPAC cohort. All panels represent the results from a simple slopes analysis. The relationship between prenatal maternal EPDS (x-axis) and offspring outcome (y-axis) is stratified by higher (+1 SD; blue line), lower (-1 SD; red line) or mean (grey line) microglial PGS in offspring. A) The microglial PGS moderates the relationship between prenatal maternal EPDS and mild depression at 24 years of age in females (N=1461; lower PGS-slope=0.02, SE=0.03, P=0.46 ; higher PGS-slope=0.11, SE=0.03, P=5.82E-5), measured by the CIS-R. B) The microglial PGS moderates the relationship between prenatal maternal EPDS and moderate depression at 24 years in females (N=1461; lower PGS-slope=0.02, SE=0.03, P=0.56 ; higher PGS-slope=0.12, SE=0.03, P=9.26E-5), measured by the CIS-R. C) The microglial PGS moderates the relationship between prenatal maternal EPDS and severe depression at 24 years in females (N=1460; lower PGS-slope=-0.03, SE=0.07, P=0.69 ; higher PGS-slope=0.18, SE=0.07, P=0.01), measured by the CIS-R.

### An adult microglial PGS moderates the association of adult risk factors on depressive symptoms in the UK Biobank

Adult exposures such as high BMI (50) or stressful life events (11) are well-characterized risk factors with evidence for depression causation. We thus asked whether an adult microglial PGS could moderate the relation between adult risk factors for depression and depressive outcomes. We created an adult microglial PGS using similar methods to the fetal microglial PGS. We identified microglial enriched genes in the subgenual anterior cingulate cortex using a dataset from Tran et al (58). This is the brain area with the strongest causal evidence for a role in depression (59). We used eQTLs from the anterior cingulate of the gene-tissue expression (GTEx) consortium to generate a PGS representative of variation in the expression of microglial enriched genes in all individuals of the UK Biobank (60). Our hypothesis required the risk factor (e.g., a stressful experience) to occur before the evaluation of depression, this precluded us from using the most common definition of depression in the UK Biobank, lifetime incidence. Instead, we used answers to the patient health questionnaire 9 (PHQ9), a measure of current depression, as our outcomes of interest. This was advantageous for two primary reasons. First, it was administered concurrently with the questionnaires querying stressful life exposures providing the required temporal ordering of events. Second, it enabled an analysis of the nine DSM symptoms of depression. We conducted multivariable regression to estimate the interaction effect between BMI or stressful life events and the adult microglial PGS with the first 40 principal components, age, assessment center attended, and genotype array used as covariates in the analysis. All analyses were stratified by sex. We used the individual answers to the nine questions of the PHQ9 and their cumulative score as continuous outcomes. There were four exposures in females with a significant interaction with the microglial PGS that passed correction for multiple comparisons (**Figure 3A**). No interaction effects passed correction for multiple comparisons in males (**Supplementary Table 14**). Of the significant female interaction effects, BMI was the exposure for the significant interaction effect on anhedonia (beta=0.001, 95%CI 0.0009 to 0.003, P=7.74E-6, FDR P value=0.004) and tiredness (beta=0.002, 95%CI 0.0008 to 0.003, P=0.001, FDR P value=0.04), and an ability to pay rent or mortgage was the exposure for the remaining two significant interaction effects on anhedonia (beta=0.01, 95%CI 0.006 to 0.016, P=7.74E-5, FDR P value=0.007) and depressed mood (beta=0.001, 95%CI 0.005 to 0.015, P=2E-4, FDR P value=0.008; **Figure 3A**).

**Figure 3.**
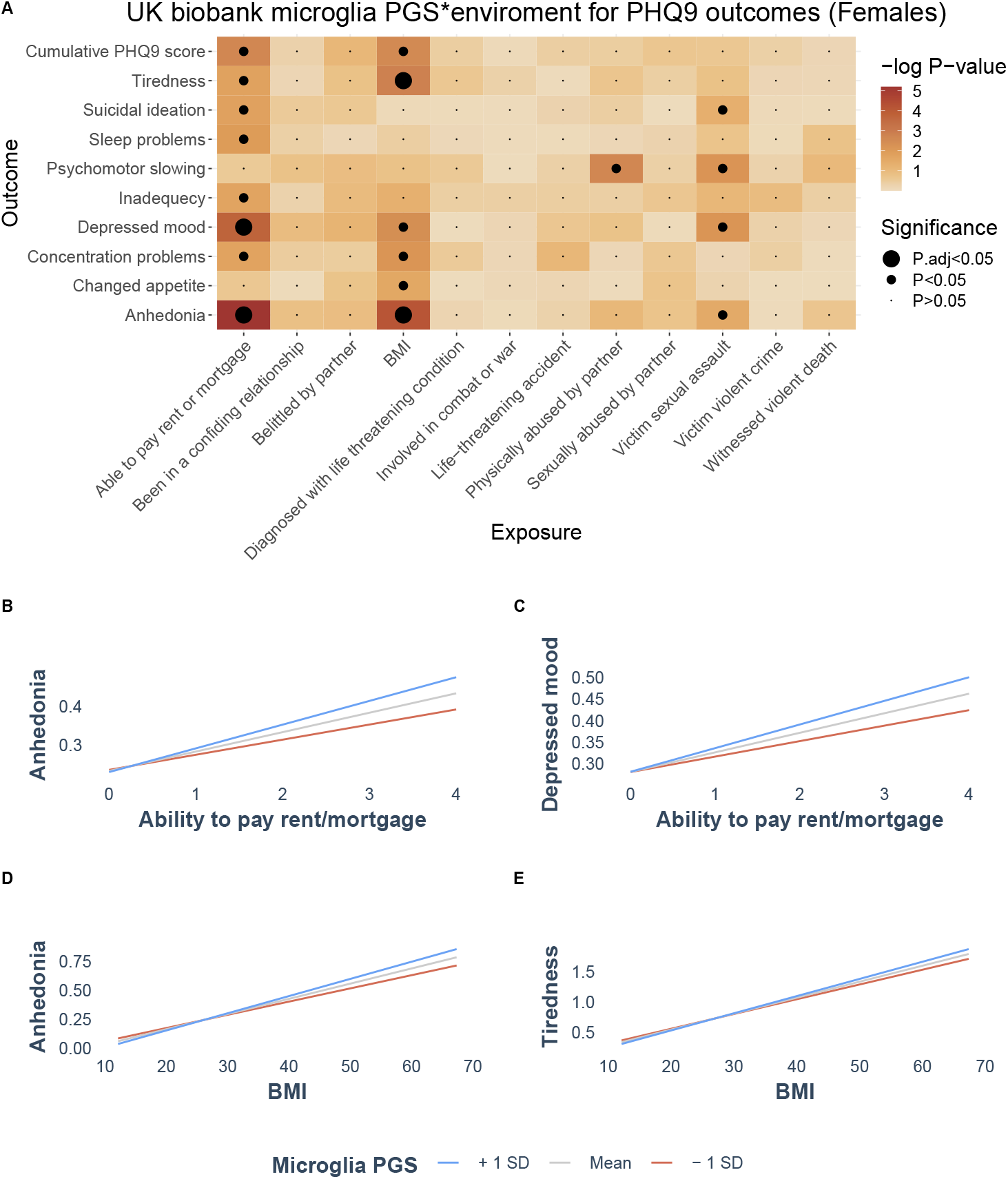
An adult microglial PGS moderates the association of risk factors on depressive symptoms in females of the UK Biobank. A) Heatmap illustrating the results from 120 linear regressions investigating the interaction between an adult microglial PGS and 12 depression risk factors (x-axis) using answers to the PHQ9 as outcomes (y-axis). Tiles are coloured by the -log of the interaction P-value, with a darker colour indicated a lower P-value. The largest dot indicates an FDR adjusted P-value < 0.05, the intermediate sized dot indicates nominal P < 0.05 and the smallest dot indicates nominal P > 0.05. There were 4 interactions that passed FDR correction. A simple slopes analysis for these interactions is plotted in B-E. In B-E, the exposure and outcome are plotted on the x- and y-axis, respectively. The relationship is stratified by higher (+1 SD; blue line), lower (-1 SD; red line) or mean (grey line) adult microglial PGS. B) Relationship between an ability to pay rent/mortgage and anhedonia in females is shown, with significant differences seen for both the higher and lower PGS slopes (N=71438; lower PGS-slope=0.04, SE=0.004, P= 6.38E-28; higher PGS-slope=0.06, SE=0.003, P=7.37E-70). C) Relationship between an ability to pay rent/mortgage and depressed mood in females is shown, with significant differences seen for both the higher and lower PGS slopes (N=71379; lower PGS-slope=0.04, SE=0.004, P= 1.58E-22; higher PGS-slope=0.05, SE=0.004, P=6.53E-70). D) Relationship between BMI and anhedonia in females is shown, with significant differences seen for both the higher and lower PGS slopes (N=72529; lower PGS-slope= 0.1, SE=0.0006, P=4.28E-76; higher PGS-slope=0.01, SE=0.0006, P=3.64E-127). E) Relationship between BMI and tiredness in females is shown, with significant differences seen for both the higher and lower PGS slopes (N=72439; lower PGS- slope=0.02, SE=0.0009, P=7.77E-164; higher PGS-slope=0.03, SE=0.0009, P=1.06E-220).

A simple slopes analysis was then conducted on the four models with a significant interaction (**Figure 3B-3E**), which showed significant associations for both the higher and lower PGS slopes in all four models (see **Figure 3B-3E** and **Supplementary Table 12**). The interpretation of these results is that a higher microglial PGS in females increases the risk of specific depressive symptoms in the presence of specific risk factors, while a lower PGS in females decreases the risk of specific depressive symptoms in the presence of specific risk factors. All associations remained significant when a depression PRS was included as a covariate (**Supplementary Table 13**). We again created PGS based on alternative cell-types for comparative purposes (astrocytes, oligodendrocyte precursor cells, excitatory neurons and inhibitory neurons using data from Tran et al). These PGS did not recapitulate the results seen in the microglial analysis, but we did observe significant interaction effects for several of the PGS that were sex- and exposure-specific (**Supplementary Figure 1**; **Supplementary Table 15 and 16**). These results describe a role for microglial function in moderating the risk of particular exposures on specific depressive symptoms in females.

## Discussion

Understanding the mechanisms of depression risk is critical for both the development of novel treatment strategies and population-based initiatives aimed at promoting mental health. We used a novel functional genomics approach in multiple independent cohorts of distinct genetic ancestries and life stages to identify microglial function as a female-specific mechanism linking adverse environmental exposures to depression. Our results identify microglial function as a potential target to mitigate the risk of depression associated with particular environmental exposures and provide a credible sex-specific mechanistic pathway to depression.

Many microglial functions are exquisitely responsive to local environmental cues(20,21,61,62), with synaptic pruning in particular garnering much attention. Compelling evidence suggests a causal role for microglial-mediated synaptic pruning in psychiatric disorders, such as schizophrenia (63,64) and studies using model systems have described sex-specific changes in microglial-mediated synaptic pruning following exposure to adverse environments (65,66). Together with these data, our results suggest further research into microglial mediated synaptic pruning, as a mechanism for sex-specific pathways in depression, is warranted. Technical and tissue limitations have limited our ability to infer such functional relations in humans, but as functional genomic resources expand, we believe studies such as ours will provide a fruitful approach for further mechanistic inference in humans.

Our findings suggest microglial function is a key regulator of depression symptom trajectory from childhood into adulthood. We identified a higher risk of internalizing and depressive symptoms in female children with a lower microglia PGS following high prenatal EPDS, but it was the higher PGS that was associated with increased depressive risk by 24 years. These findings echo previous studies, which have described multiple sex-dependent trajectories of depressive symptoms from childhood to adulthood (57,67,68). We suggest two possible mechanisms that may underlie the shift in direction of association we observe. First, adolescence is a highly dynamic period of brain maturation and microglia, through functions such as synaptic pruning, play a central role in this dynamic maturation. Individual differences in this maturation may be captured by our PGS and the trajectory of depressive symptoms may shift accordingly. Indeed shifts in developmental trajectories are commonly hypothesized to underlie the relationship between early life events and psychopathologies (69–71). The second, and perhaps more parsimonious, explanation is differences in sex hormones from childhood to adulthood, which differentially engage with either microglia or neuronal elements previously shaped by microglial function. There has been little research into the relation between microglia and sex hormones during adolescence and future work evaluating the evolution of depressive symptoms in longitudinal cohorts stratified by both sex and adverse exposures will be highly informative.

Microglial function is often defined by interaction with other cell-types, for instance synaptic pruning and effects on myelination. In both GUSTO and ALSPAC we did not identify any associations using alternative cell-type PGS, but in the UK Biobank we observed striking cell and sex-specific associations. These associations are likely a function of both the granular phenotype information available and the large sample size of the UK Biobank. The etiology of depression risk is extraordinarily complex, and this complexity certainly involves the interaction between many cell-types. Our results are consistent with this heterogeneity but implicate microglia as a negotiator central in these interactions.

The role of microglia in depression risk and progression is becoming more widely appreciated (72). There is a huge demand for novel anti-depressants, particularly those that target alternative pathways to traditional anti-depressants, as they may be of use for individuals resistant to current approaches. Our work suggests designing therapeutic strategies around microglial homeostatic function may be a novel therapeutic approach.

It should be noted that our study also has several limitations. Although our analysis contained several genetic ancestries, we were not able to sufficiently sample all populations. Emerging biobanks, particularly in Africa, will be of huge importance in advancing equitable depression research. Our study also only included males and females that identified with their sex at birth.

Throughout our study we used cis-eQTLs, these variants do not fully explain the genomic contribution to variation in gene expression and as functional genomic resources expand, the discovery power of studies such as ours will increase enormously. Analysis in GTEx show few sex-biased brain eQTLs (73), therefore we do not expect sex differences in the genomic architecture of gene expression to significantly bias our findings. However, with the increasing sample size of functional genomics resources, using sex-specific functional variants in future studies will be an interesting avenue.

Our study provides a framework to assess cell-specific involvement in human health and disease. We describe microglia as a critical player in the sex-specific translation of environmental exposures into depression risk. Our findings have direct implications for the understanding of the sex-biases in depression prevalence.

## Data Availability

All data used in this manuscript are available dependent on successful application to the relevant cohort study

## Author contributions

EF planned the study, conducted the analysis, interpreted the data, and wrote the manuscript, with editing from MJM. IP and SP contributed to the analysis and genetic scores. SYC, APT, PPS and MJM aided in study design. All authors approved the final version of the manuscript.

## Acknowledgments

We would like to acknowledge both the GUSTO, ALSPAC and UK Biobank investigators, staff, researchers and in particular the participants. This research has been conducted using the UK Biobank Resource under Application Number 41975.

The GUSTO study is supported by grants NMRC/TCR/004-NUS/2008 and NMRC/TCR/012-NUHS/2014 from the Singapore National Research Foundation under the Translational and Clinical Research Flagship and grant OFLCG/MOH-000504 from the Open Fund Large Collaborative Grant Programmes and administered by the Singapore Ministry of Health’s National Medical Research Council (NMRC), Singapore.

## Data availability

Access to data from the GUSTO, ALSPAC and UK Biobank are dependent on approved application to the respective data access committees. All other data generated in this study are provided in the supplementary material.

We are extremely grateful to all the families who took part in this study, the midwives for their help in recruiting them, and the whole ALSPAC team, which includes interviewers, computer and laboratory technicians, clerical workers, research scientists, volunteers, managers, receptionists and nurses. The UK Medical Research Council and Wellcome (Grant ref: 217065/Z/19/Z) and the University of Bristol provide core support for ALSPAC. A comprehensive list of grants funding is available on the ALSPAC website: http://www.bristol.ac.uk/alspac/external/documents/grant-acknowledgements.pdf.

The Genotype-Tissue Expression (GTEx) Project was supported by the Common Fund of the Office of the Director of the National Institutes of Health, and by NCI, NHGRI, NHLBI, NIDA, NIMH, and NINDS. The data used for the adult eQTL analyses described in this manuscript were obtained from the GTEx Portal.

## Figure captions

**Supplementary figure 1**

Heatmap illustrating the results from linear regressions investigating the interaction between 4 alternative cell-type PGS (astrocyte, excitatory neuron, inhibitory neuron and oligodendrocyte precursor cell (OPC)) and 12 depression risk factors (x-axis) using answers to the PHQ9 as outcomes (y-axis). Tiles are coloured by the -log of the interaction P-value, with a darker colour indicated a lower P-value. The largest dot indicates an FDR adjusted P-value < 0.05, the intermediate sized dot indicates nominal P < 0.05 and the smallest dot indicates nominal P > 0.05. Results are shown separately for females (A) and males (B).

