## Supplementary figures and images for "Microglial function moderates the relation between depression risk factors and depression outcomes across the life course in females"

### Supplemental figure

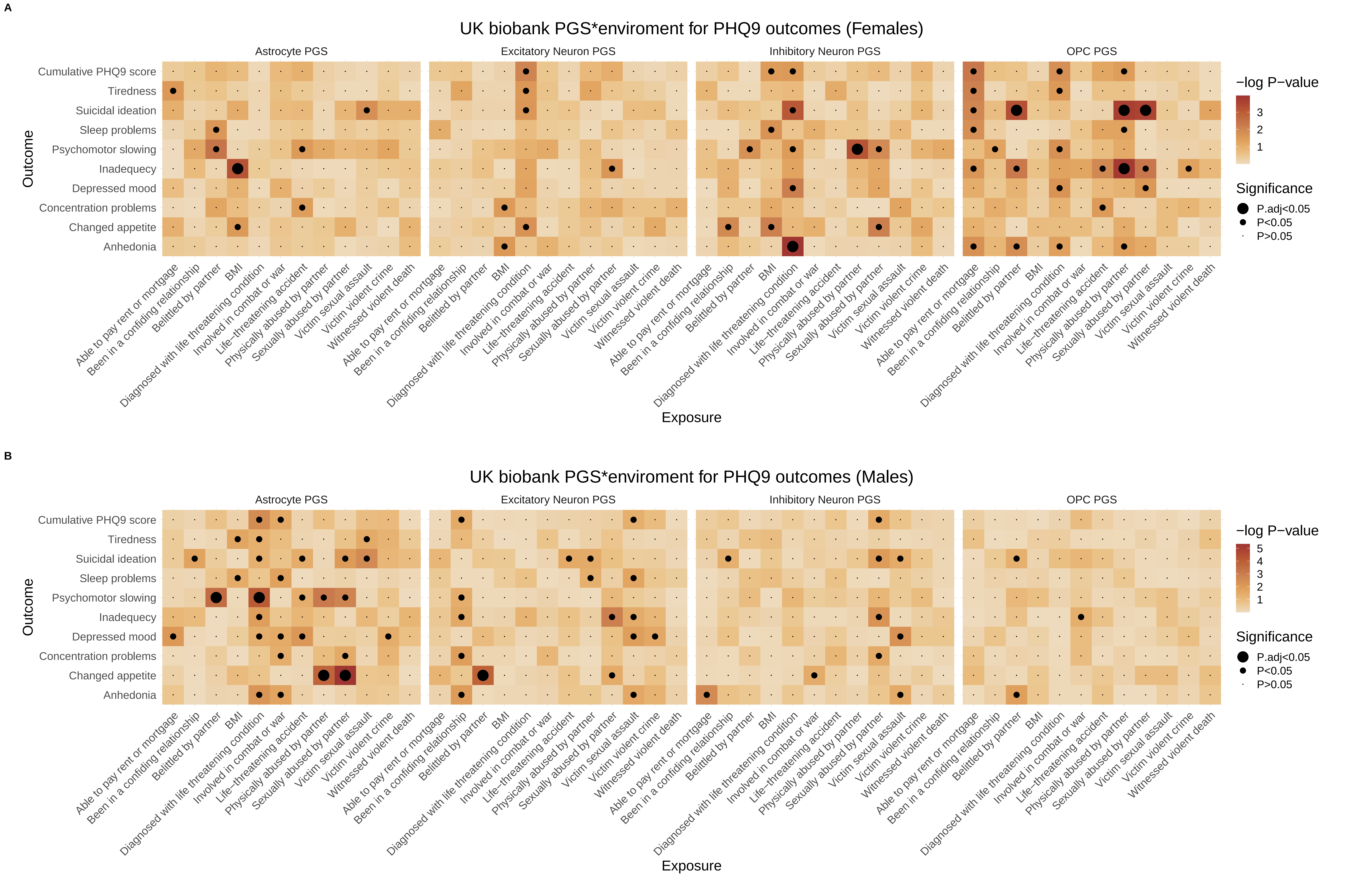
